# Metabolic dysfunction-associated steatotic liver disease status modifies risks of in-stent restenosis in coronary atherosclerosis: A long-term longitudinal study

**DOI:** 10.1101/2025.02.19.25322577

**Authors:** Jiaming Lai, Junzhao Ye, Ling Luo, Long Teng, Congxiang Shao, Zhi Dong, Weiyi Mai, Xiaodong Zhuang, Bihui Zhong

## Abstract

**Background:** Previous studies have verified that metabolic dysfunction-associated steatotic liver disease (MASLD) confered higher risk of coronary atherosclerosis development. However, whether MASLD influence prognosis after drug-eluting stent (DES) implantation treatment remain not known.

**Methods:** In this retrospective observational study, 301 included cardiovascular disease (CVD) patients who underwent re-coronary angiography after the first successful DES-based percutaneous coronary intervention. All the patients received computerized tomography (CT) to estimate liver steatosis (65.8% of MASLD). The primary outcome was in-stent restenosis (ISR) determined by intravenous ultrasound. Liver fibrosis was assessed with Fibrosis-4 (FIB-4) index.

**Results:** After a median follow-up of 27 (range from 12 to 144) months, subjects with MASLD over presented ISR than those without (30.3% vs. 8.7 %, P < 0.001). The Cox proportional hazard model confirmed that, MASLD [HR (95%CI): 2.64 (1.14– 6.11), P = 0.024], FIB-4 index [HR (95%CI): 2.05 (1.50–2.82), P < 0.001] were independently associated with ISR. The hazard model’s area under the receiver operating characteristic curves (AUROC) of 1, 3, 5 and 10 years prediction for ISR were respectively 0.620, 0.801, 0.830 and 0.721. Kaplan-Meier survival analysis demonstrated that ISR increased progressively with the FIB-4 index (log-rank, P<0.001). Additionally, after low-density lipoprotein (LDL) cholesterol reached control standard, FIB-4 index [HR (95%CI): 2.72 (1.43–5.16), P = 0.002] and liver CT attenuation [HR (95%CI): 0.94 (0.88–0.99), P = 0.048] remained independently associated with ISR.

**Conclusions:** MASLD and related liver fibrosis are associated with the ISR in CVD patients after DES implantation, and management of MASLD might attenuate the risks of ISR.

**Graphical abstract:** 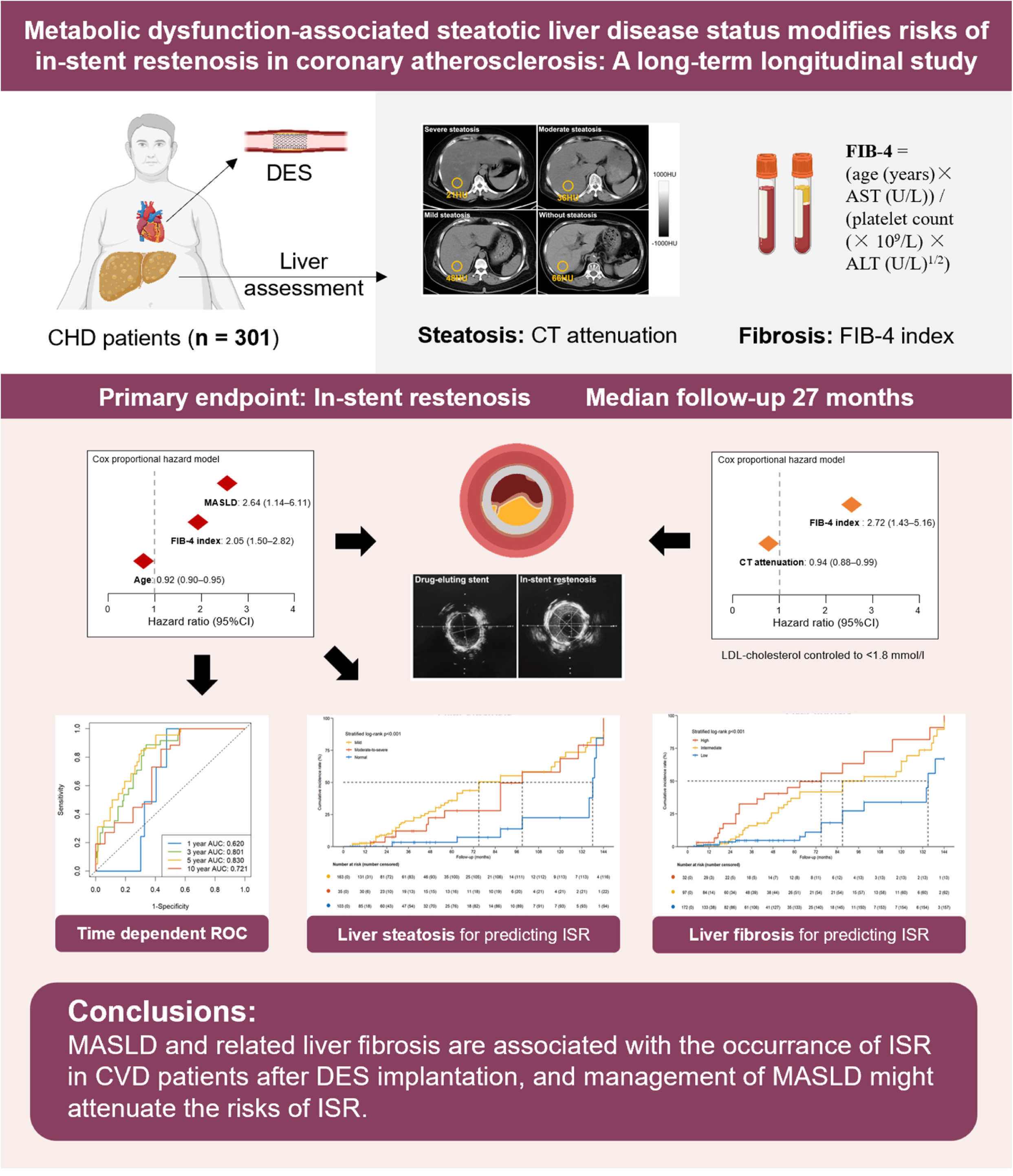

## Introduction

Metabolic dysfunction-associated steatotic liver disease (MASLD) has been recognized as a strikingly increasing source of chronic liver disease burden globally, with an estimated prevalence of 30% [1,2]. MASLD progression and related severity has a bidirectional association with cardiovascular disease (CVD) and its risk factors encompassing metabolic syndrome, and type 2 diabetes [3]. Of notes, CVD accounts for the leading cause of death in MASLD patients and MASLD has been ascribed as emerging drivers of CVD, A meta-analysis involved 67,070 adult patients with MASLD showed that the combined prevalence of coronary heart disease (CHD) was 44.6% (95% confidence interval (CI): 36.0%-53.6%) [4]. However, whether MASLD have an impact on CVD after treatment remains undetermined.

Percutaneous coronary intervention (PCI) is the primary treatment strategy for CVD, while in-stent restenosis (ISR) following PCI refers to the narrowing of the stent lumen or 5 mm segments at both ends of the stent detected by coronary angiography, resulting in a stenosis degree ≥ 50% [5]. Although drug-eluting stents (DES) significantly lower the incidence of ISR by diminishing the local inflammatory response and inhibiting arterial intimal hyperplasia through drug release, the occurrence rate of ISR ranges from 3 to 20% [6] and become crucial challenges in the prognosis of CVD. To our knowledge, only one research with ultrasound grapy assessments of steatosis implicated that MASLD might elevate risk of in-stent restenosis after bare metal stenting, whereas whether it also affect treatment outcomes of those received drug-eluting stent implantation remained not known, of which prevent restenosis that may arise with traditional metal stents via gradually releasing the drug in the coating to inhibit the excessive proliferation of vascular smooth muscle cells [7].

In this era of DES implantation during PCI, the occurrence of ISR after DES is not negligible. Therefore, we intended to investigate the role of MASLD and related liver steatosis and fibrosis in predicting ISR in patients with CHD after DES-based PCI.

## Methods

### Study design and participants

This retrospective observational cohort study was conducted utilizing data from the electronic specialize disease of Cardiovascular disease Database (CDD) managed by the at the Fatty Liver Disease Center and Cardiology of the First Affiliated Hospital of Sun Yat-sen University (Guangzhou, China). CDD is a comprehensive electronic database collecting demographic data, laboratory test information, diagnoses, treatment prescriptions, follow-up status, as well as through capturing inpatient data of the First Affiliated Hospital, Sun Yat-sen university in Guangzhou, representing the largest hospital in southern China. The study was approved by the Ethics Committee of the First Affiliated Hospital, Sun Yat-sen university {[2014]112}. All study participants provided signed informed consent. The study protocol was registered in 2014 with the definition of NAFLD and the collected variables encompassed all indicators necessary for diagnosing MASLD. In the final analysis, the patients included in this study were chosen according to the most recent diagnostic criteria for MASLD, which includes the presence of hepatic steatosis proved by imaging and at least one cardiometabolic risk factor [1].

Between January 2012 and January 2024, 1901 CHD patients, were consecutively enrolled after intravenous ultrasound (IVUS) -guided PCI admitted to the First Affiliated Hospital of Sun Yat-sen University. All patients underwent successful PCI for the culprit lesion, involving intraoperative DES implantation in accordance with the Chinese Guidelines for PCI (2016). We excluded patients with excessive alcohol intake (>20 g per day in men and 10 g per day in women), and secondary causes of steatotic liver disease (eg, chronic use of systemic corticosteroids or methotrexate). Patients with positive hepatitis B surface antigen, antibody against hepatitis C virus and antinuclear antibody titre >1/160 were also excluded. Patients were regularly followed up with PCI reassessments every 3 years or until an incident suspected symptom of chest pain, last follow-up date (144 months), depended on whichever came first. Among the 1080 CHD patients undergoing DES implantation, 423 had undergone regular follow-up PCI and IVUS. Eventually, a total of 301 patients (239 male, 63.09±10.87 years old) included in this study according to the exclusion criteria.

Moreover, LDL-cholesterol is the primary lipid-lowering target for CVD. We found that patients who achieved LDL-cholesterol control standard may still develop ISR. Therefore, we also collected the data of LDL-cholesterol of patients who were regularly followed up at the last follow-up date. According to 2024 Chinese guidelines for the diagnosis and management of patients with chronic coronary syndrome, LDL-cholesterol control standard was defined as <1.8 mmol/L and decreased by >50% from baseline, by using lipid-lowering drugs such as statins or lifestyle therapies [8].

### Clinical information

For clinical data extracted from CDD database, sociodemographic characteristics such as age, sex, diabetes mellitus, hypertension, smoking history, and drinking history, along with clinical presentation, were documented. Anthropometric measurements including body weight, body height and waist circumference were captured. Body mass index (BMI) was calculated as weight (kg) divided by height (m) squared. Laboratory analyses included measurements of fasting glucose; lipid profile (total cholestero, triglycerides, high- and low-density lipoprotein cholesterol (HDL and LDL, respectively), Lp(a) and other apolipoproteins,); kidney function (blood urea nitrogen, creatinine and urate) levels) and liver function (aspartate transaminase (AST), alanine aminotransferase (ALT), alkaline phosphate, and gamma-glutamyltransferase; and serum albumin. Furthermore, the utilization of pharmaceutical agents such as aspirin, clopidogrel, statins, beta blockers, and angiotensin-converting enzyme inhibitor or angiotensin receptor blocker was recorded.

### Liver assessments

Liver steatosis was measured by computerized tomography (CT) by at least two experienced radiologists. CT images were acquired with a SOMATOM Force CT Scanner (Siemens Medical Solutions, Munich, Germany). An abdominal non-contrast CT scan was performed just before the PCI. Hepatic and splenic Hounsfield units (HU) were measured in the largest possible regions. The regions of interest >100 mm^2^ included two areas that were aligned with the anterior-posterior dimension of the right liver lobe and one that was aligned with the spleen. In the liver, we located regions of interest by avoiding the inclusion of any large vessels or biliary structures. A liver-to-spleen attenuation ratio of <1.0 was defined as the cut-off for a positive diagnosis of mild liver steatosis and a hepatic attenuation of < 40 HU corresponds to a moderate-to-severe steatosis [9]. Representative liver CT images are shown in Figure 1C.

**Figure 1.**
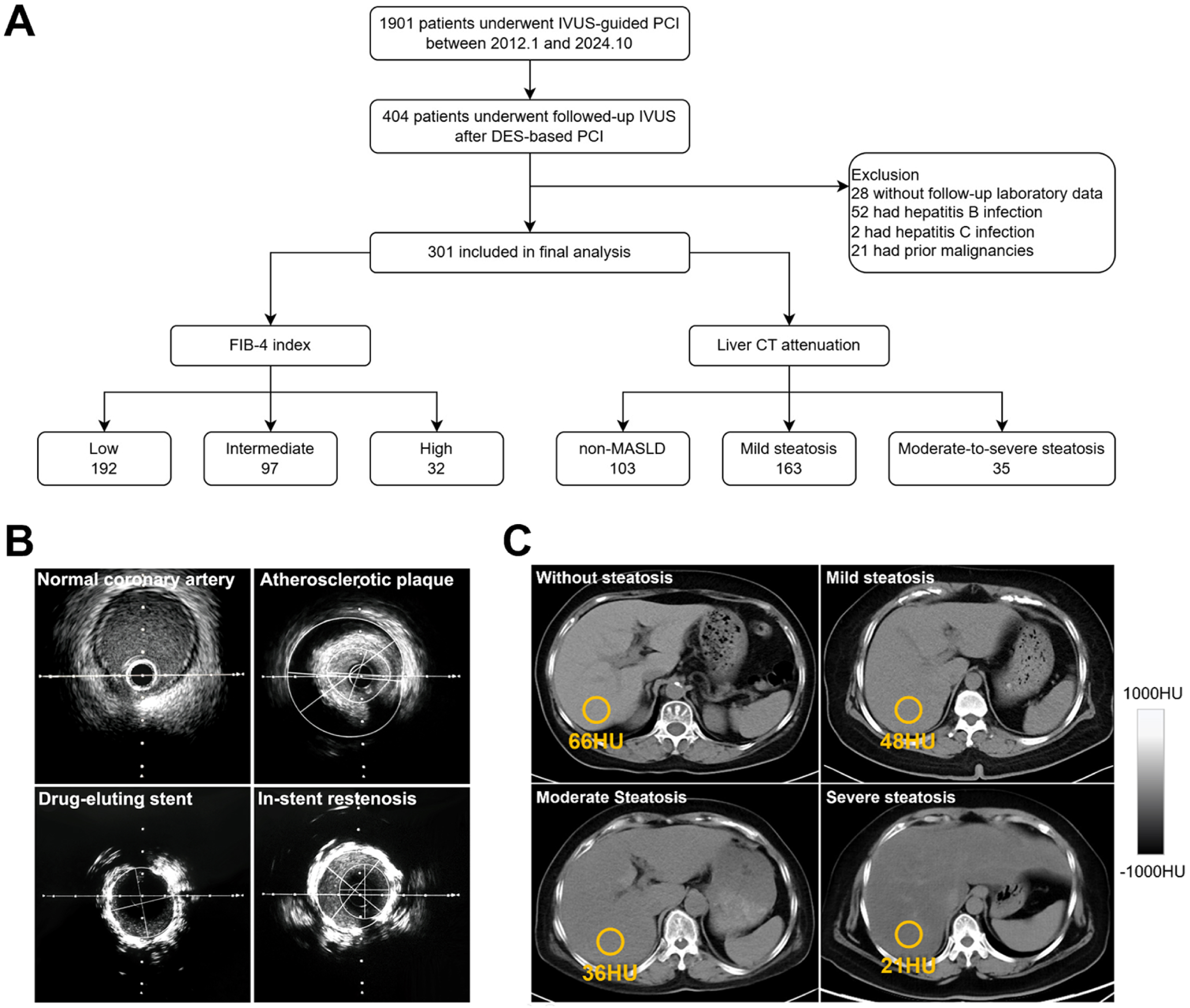
**A)** Flowchart of the patient selection process. **B)** Representative intravenous ultrasound images: Normal coronary artery, atherosclerotic plaque, drug-eluting stent and In-stent restenosis respectively. **C)** Representative liver CT images quantified liver steatosis using the absolute unenhanced CT attenuation. Normal liver without steatosis (> 57 HU), mild steatosis (40–57 HU), moderate steatosis (23–40 HU) and severe steatosis (< 23 HU) respectively. **Abbreviations:** IVUS: intravenous ultrasound; PCI: percutaneous coronary intervention; DES: drug-eluting stent; MASLD: metabolic dysfunction-associated steatotic liver disease; CT: computerized tomography; HU: hounsfield units.

For liver fibrosis, fibrosis-4 index (FIB-4) was use as a common noninvasive measurement of liver fibrosis for its simplicity and accessibility. FIB-4 = (age (years)× AST (U/L)) / (platelet count (× 10^9^/L) × ALT (U/L)^1/2^). Based on the EASL MASLD guideline, we classified the values of the FIB-4 index as follows: low group (<1.3 or <2.0 in individuals aged >65), intermediate group (1.30-2.67 or 2.0-2,67 in individuals aged >65), and high group (>2.67 for all ages) [1].

### Intravenous ultrasound characteristics and outcome

The diagnosis of coronary artery disease was based on cardiac catheterisation findings that were reviewed by at least two experienced cardiologists. Quantitative IVUS analysis was performed using the built-in iMap planimetry function in iMap-IVUS (Boston Scientific, Natick, MA, USA). Initially, areas of lumen and vessel inside an external elastic membrane were manually traced to determine the minimum lumen area at every 1-mm interval in diseased segments. Thereafter, areas of atherosclerotic plaque were calculated at the most severe stegnotic lesion. Significant coronary artery disease was defined as the presence of at least 50% stenosis at one or more major coronary arteries. After the procedure, medical therapy, percutaneous coronary intervention and coronary artery bypass grafting were provided as clinically indicated. Upon discharge, the patients were followed up at the cardiac clinic every 3 to 6 months. During each visit, patients were reviewed for recurrence of angina, functional status, blood pressure and drug regime. ISR following IVUS refers to the narrowing of the stent lumen or 5 mm segments at both ends of the stent detected by coronary angiography, resulting in a stenosis degree ≥ 50%. When ISR was diagnosed by the follow-up angiography and IVUS during the study period, patients were treated with re-DES implantation. The successful procedure was defined as a reduction of the stenosis to <10% residual narrowing, thrombolysis in myocardial infarction flow Grade III, with improvement in ischemic symptoms, and without major procedure related complications [5]. Representative IVUS images are shown in Figure 1B.

### Statistical analysis

In the current study, the database was frozen when the last recruited patient had reached 1 year of follow-up. Continuous variables were expressed as mean±SD or median (IQR), and compared between patients with and without MASLD using unpaired t test or ManneWhitney U test as appropriate. Categorical variables were compared using c^2^ test. Binary logistic regression analysis was performed to identify independent factors associated with coronary artery disease. Time-to-event curves of patients with and without MASLD were compared using the log-rank test. The Cox proportional hazard model was used to identify independent factors associated with the ISR. Factors with p values <0.05 by univariate analysis were included in the multivariate analysis. We further used a receiver operating characteristic (ROC) curve and the areas under the receiver operator characteristic curve (AUC) to evaluate the efficiency of MASLD related factors in predicting ISR. All statistical analysis was performed using SPSS version 16.0 and R version 4.3.3. Statistical significance was taken as a two-sided p value of <0.05.

## Results

### Clinical Characteristics

423 regularly followed up patients were screened for eligibility. After excluding patients without laboratory data, those with viral hepatitis and prior malignancies, 301 patients were enrolled in the current study (Figure 1A). Overall, 198 (65.8%) patients had MASLD. No significant differences were observed between age, genders, BMI, risk factors for CHD (including current smoking, drinking, diabetes and hypertension) (all P>0.05). However, statistically significant differences as higher in the total cholesterol, triglyceride, fasting glucose, uric acid, alanine aminotransferase, aspartate aminotransferase, γ-glutamyl transpeptidase and liver computerized tomography attenuation were observed in MASLD patients that a (all P<0.05). Additionally, MASLD patients had higher application rate of stains (65.15% vs 42.72%, P<0.001) as presented in Table 1.

**Table 1.** Clinical baseline characteristics of patients.

| Characteristics | Total<br>(n = 301) | MASLD<br>(n = 198) | Non MASLD<br>(n = 103) | *p* value |
| --- | --- | --- | --- | --- |
| Age (years) | 63.09 ± 10.87 | 62.65 ± 11.32 | 63.93 ± 9.96 | 0.333 |
| Male, n (%) | 239 (79.40%) | 155 (78.28%) | 84 (81.55%) | 0.506 |
| Smoking, n (%) | 145 (48.17%) | 96 (48.48%) | 49 (47.57%) | 0.881 |
| Alcohol, n (%) | 79 (26.42%) | 55 (27.78%) | 24 (23.76%) | 0.456 |
| Diabetes, n (%) | 130 (43.19%) | 90 (45.45%) | 40 (38.83%) | 0.271 |
| Hypertension, n (%) | 211 (70.10%) | 143 (72.22%) | 68 (66.02%) | 0.265 |
| Systolic blood pressure (mm Hg) | 130.91 ± 17.95 | 129.16 ± 18.22 | 134.29 ± 16.99 | 0.060 |
| Diastolic blood pressure (mm Hg) | 76.11 ± 11.28 | 76.29 ± 11.66 | 75.76 ± 10.56 | 0.697 |
| Body mass index (kg/m<sup>2</sup>) | 26.10 ± 3.09 | 26.13 ± 3.48 | 26.04 ± 2.24 | 0.782 |
| Waist circumference (cm) | 91.77 ± 7.48 | 92.00 ± 8.03 | 91.22 ± 5.97 | 0.437 |
| Total cholesterol (mmol/L) | 4.45 ± 1.41 | 4.56 ± 1.36 | 4.22 ± 1.48 | **0.044** |
| Triglyceride (mmol/L) | 1.55 (1.10, 2.16) | 1.61 (1.13, 2.29) | 1.43 (1.04, 1.89) | **0.019** |
| HDL cholesterol (mmol/L) | 0.98 ± 0.24 | 1.00 ± 0.24 | 0.96 ± 0.23 | 0.159 |
| LDL cholesterol (mmol/L) | 2.81 ± 1.03 | 2.87 ± 0.99 | 2.69 ± 1.09 | 0.166 |
| ApoA1 (g/L) | 1.11 ± 0.23 | 1.13 ± 0.24 | 1.06 ± 0.19 | **0.022** |
| ApoB (g/L) | 0.76 ± 0.30 | 0.77 ± 0.29 | 0.76 ± 0.31 | 0.876 |
| ApoE (mg/L) | 36.80 ± 17.88 | 38.62 ± 20.17 | 33.58 ± 12.28 | **0.011** |
| Lp(a) (mg/L) | 174.00 (79.00, 415.00) | 166.00 (78.25, 388.00) | 181.00 (82.00, 443.00) | 0.350 |
| Fasting glucose (mmol/L) | 6.25 (5.20, 8.10) | 5.85 (5.10, 7.38) | 6.50 (5.40, 8.57) | **0.026** |
| Hemoglobin A1c, (%) | 6.85 ± 1.68 | 6.95 ± 1.69 | 6.67 ± 1.66 | 0.172 |
| Uric acid (μmol/L) | 414.73 ± 116.67 | 427.95 ± 116.09 | 388.28 ± 113.85 | **0.006** |
| Urea nitrogen (mmol/l) | 5.70 (4.80, 7.40) | 5.60 (4.80, 7.40) | 5.80 (4.80, 7.30) | 0.868 |
| Creatinine (mmol/l) | 84.00 (70.00, 98.75) | 85.00 (70.00, 99.00) | 81.00 (71.00, 98.00) | 0.544 |
| Alanine aminotransferase (U/L) | 28.00 (20.00, 38.00) | 31.00 (22.00, 41.00) | 26.00 (17.50, 32.00) | **<0.001** |
| Aspartate aminotransferase (U/L) | 26.00 (22.00, 34.00) | 29.00 (23.00, 35.75) | 23.00 (20.00, 28.50) | **<0.001** |
| γ-glutamyl transpeptidase (U/L) | 35.00 (24.00, 55.00) | 38.00 (27.00, 56.00) | 28.50 (21.00, 45.00) | **0.002** |
| Albumin (g/L) | 40.39 ± 3.83 | 40.63 ± 3.89 | 39.92 ± 3.69 | 0.129 |
| Platelet (10<sup>9</sup>/L) | 223.23 ± 59.04 | 221.35 ± 60.20 | 226.83 ± 56.85 | 0.446 |
| Liver CT attenuation (HU) | 56.00 (51.00, 63.00) | 54.00 (48.50, 56.00) | 67.00 (62.50, 70.00) | **<0.001** |
| FIB-4 index | 1.51 (1.15, 1.88) | 1.54 (1.18, 1.89) | 1.39 (1.06, 1.73) | 0.079 |
| Drugs, n (%) |  |  |  |  |
| Aspirin | 272 (90.37%) | 176 (88.89%) | 96 (93.20%) | 0.229 |
| Clopidogrel | 276 (91.69%) | 179 (90.40%) | 97 (94.17%) | 0.261 |
| Antihypertension | 224 (74.42%) | 154 (77.78%) | 70 (67.96%) | 0.064 |
| Hypoglycemics | 134 (44.52%) | 94 (47.47%) | 40 (38.83%) | 0.152 |
| Stains | 173 (57.48%) | 129 (65.15%) | 44 (42.72%) | **<0.001** |
| Indication of coronary angiogram, n (%) |  |  |  | 0.125 |
| Myocardial infarction | 59 (19.60%) | 44 (22.22%) | 15 (14.56%) |  |
| Unstable angina | 198 (65.78%) | 126 (63.64%) | 72 (69.90%) |  |
| Stable coronary heart disease | 34 (11.30%) | 24 (12.12%) | 10 (9.71%) |  |
| Others | 10 (3.32%) | 4 (2.02%) | 6 (5.83%) |  |
| Follow-up duration (months) | 27.00 (15.00, 56.00) | 27.00 (15.00, 56.00) | 27.00 (15.00, 51.00) | 0.848 |

Continuous variables were expressed as mean ± SD or median (IQR).
HDL, high-density lipoprotein; LDL, low-density lipoprotein; CT, computerized tomography.

### The relationship between ISR development and MASLD

To evaluate the interaction between MASLD and ISR, we included 301 patients with CHD, at a mean follow-up of 27 (range from 12 to 144) months, 60 of 198 (30.3%) patients with MASLD and 9 of 103 (8.7%) patients without MASLD reached the end point of ISR [HR (95%CI): 3.522(1.746–7.105), P<0.001 by log-rank test].

We conducted Cox proportional hazard model to find out the relationship between ISR development and MASLD. By univariate analysis MASLD, age, female gender, fasting glucose, total cholesterol, triglycerides, FIB-4 index and liver computerized tomography attenuation were associated with ISR (all P < 0.05). However, LDL-cholesterol and Hemoglobin A1c showed a relative relation with ISR with borderline statistical significance (P = 0.059 and P = 0.050). By multivariate analysis, MASLD, FIB-4 index and age were independent factors associated with the end point. Additionally, MASLD [HR (95%CI): 2.64 (1.14–6.11), P = 0.024] and FIB-4 index [HR (95%CI): 1.98 (1.44 ∼ 2.73), P<0.001] had a positive association with ISR, while age had a negative association with ISR [HR (95%CI): 0.93 (0.90– 0.95), P < 0.001] (Table 2).

**Table 2.** Factors associated with in-stent restenosis in patients with coronary heart disease.

| Factors | Univariate analysis |  | Multivariate analysis |  |
| --- | --- | --- | --- | --- |
|  | HR (95% CI) | *p* Value | HR (95% CI) | *p* Value |
| MASLD | 3.49 (1.73 ~ 7.03) | **<0.001** | 2.64 (1.14 ~ 6.11) | **0.024** |
| Age (years) | 0.95 (0.93 ~ 0.98) | **<0.001** | 0.92 (0.90 ~ 0.95) | **<0.001** |
| Female gender | 1.77 (1.01 ~ 3.09) | **0.044** | 1.79 (0.95 ~ 3.37) | 0.073 |
| Smoking | 1.01 (0.62 ~ 1.65) | 0.967 |  |  |
| Alcohol | 1.23 (0.70 ~ 2.15) | 0.479 |  |  |
| Diabetes | 1.49 (0.88 ~ 2.54) | 0.142 |  |  |
| Hypertension | 0.73 (0.41 ~ 1.28) | 0.269 |  |  |
| Systolic blood pressure (mm Hg) | 0.99 (0.98 ~ 1.01) | 0.219 |  |  |
| Diastolic blood pressure (mm Hg) | 1.02 (1.00 ~ 1.04) | 0.088 |  |  |
| Body mass index (kg/m<sup>2</sup>) | 1.05 (0.96 ~ 1.15) | 0.310 |  |  |
| Waist circumference (cm) | 1.03 (0.99 ~ 1.07) | 0.167 |  |  |
| Fasting glucose (mmol/l) | 1.06 (1.01 ~ 1.11) | **0.035** | 1.04 (0.97 ~ 1.12) | 0.251 |
| Total cholesterol (mmol/l) | 1.20 (1.02 ~ 1.42) | **0.031** | 0.97 (0.80 ~ 1.18) | 0.765 |
| Triglycerides (mmol/l) | 1.48 (1.18 ~ 1.87) | **<0.001** | 1.16 (0.85 ~ 1.58) | 0.359 |
| HDL-cholesterol (mmol/l) | 0.80 (0.31 ~ 2.08) | 0.652 |  |  |
| LDL-cholesterol (mmol/l) | 1.26 (0.99 ~ 1.60) | **0.059** |  |  |
| ApoA1 (g/L) | 0.80 (0.23 ~ 2.78) | 0.720 |  |  |
| ApoB (g/L) | 0.85 (0.34 ~ 2.16) | 0.734 |  |  |
| ApoE (mg/L) | 1.01 (0.99 ~ 1.02) | 0.212 |  |  |
| Lp-a (mg/L) | 1.00 (1.00 ~ 1.00) | 0.787 |  |  |
| Uric acid (μmol/L) | 1.00 (1.00 ~ 1.00) | 0.540 |  |  |
| Creatinine (mmol/l) | 1.00 (0.99 ~ 1.00) | 0.218 |  |  |
| Alanine aminotransferase (U/L) | 1.00 (0.98 ~ 1.01) | 0.689 |  |  |
| Platelet (10<sup>9</sup>/L) | 1.00 (1.00 ~ 1.00) | 0.920 |  |  |
| Hemoglobin A1c (%) | 1.20 (1.00 ~ 1.45) | **0.050** |  |  |
| FIB-4 index | 1.52 (1.13 ~ 2.04) | **0.002** | 2.05 (1.50 ~ 2.82) | **<0.001** |
| Liver CT attenuation (HU) | 0.96 (0.94 ~ 0.98) | **<0.001** | 1.00 (0.96 ~ 1.03) | 0.699 |
| Drug use |  |  |  |  |
| Aspirin | 0.95 (0.43 ~ 2.07) | 0.891 |  |  |
| Clopidogrel | 1.49 (0.66 ~ 3.33) | 0.335 |  |  |
| Antihypertension | 0.68 (0.38 ~ 1.23) | 0.201 |  |  |
| Hypoglycemics | 1.38 (0.85 ~ 2.24) | 0.197 |  |  |
| Stains | 1.05 (0.63 ~ 1.75) | 0.841 |  |  |

HDL, high-density lipoprotein; LDL, low-density lipoprotein; CT, computerized tomography.

We incorporated these 3 variables found by multivariate cox regression analysis (MASLD, FIB-4 index and age) into the risk prediction model and conducted time dependent ROC analysis on it. The result showed that the AUC of 1, 3, 5 and 10 years prediction for ISR were respectively 0.620 (95%CI: 0.532–0.709), 0.801 (95%CI: 0.721–0.880), 0.830 (95%CI: 0.755–0.905) and 0.721 (95%CI: 0.561–0.881). The AUC reached above 0.8 for 3 and 5 years prediction, which showed that this model had good predictive value for ISR (Figure 3A).

### The relationship between ISR and the severity of MASLD

Patients with different liver fibrosis stage were divided into three groups according to FIB-4 index. The median values of the groups with low, intermediate, and high FIB-4 index were 1.19 [0.95–1.54, n=172 (57.1%)], 1.69 [1.51–2.06, n=97 (32.2%)], and 2.90 [2.76–3.16, n=32 (10.6%)], respectively (P < 0.001). Kaplan-Meier survival analysis showed that ISR increased progressively with the high and intermediate FIB-4 index group, the median survival time were 75 and 87 months, while the median survival time of low FIB-4 index group is 135 months (stratified log-rank, P < 0.001) (Figure 2A). In these three FIB-4 index groups, the respective 1, 3, 5, and 10 year cumulative incidence rates of ISR were 1.2% (95% CI: 0.0–2.8), 2.9% (95% CI: 0.4–5.4), 2.9% (95% CI: 0.4–5.4), and 7.0% (95% CI: 3.2–10.8) in the low FIB-4 index group; 1.0% (95% CI: 0.0–3.0), 10.3% (95% CI: 4.3–16.4), 20.6% (95% CI: 12.6–28.7), and 27.8% (95% CI:18.9–36.8) in the intermediate FIB-4 index group; and 3.1% (95% CI: 0.0–9.2), 28.1% (95% CI: 12.5–43.7), 37.5% (95% CI: 20.7–54.3), and 53.1% (95% CI: 35.8–70.4) in the high FIB-4 index group (Figure 2B).

**Figure 2.**
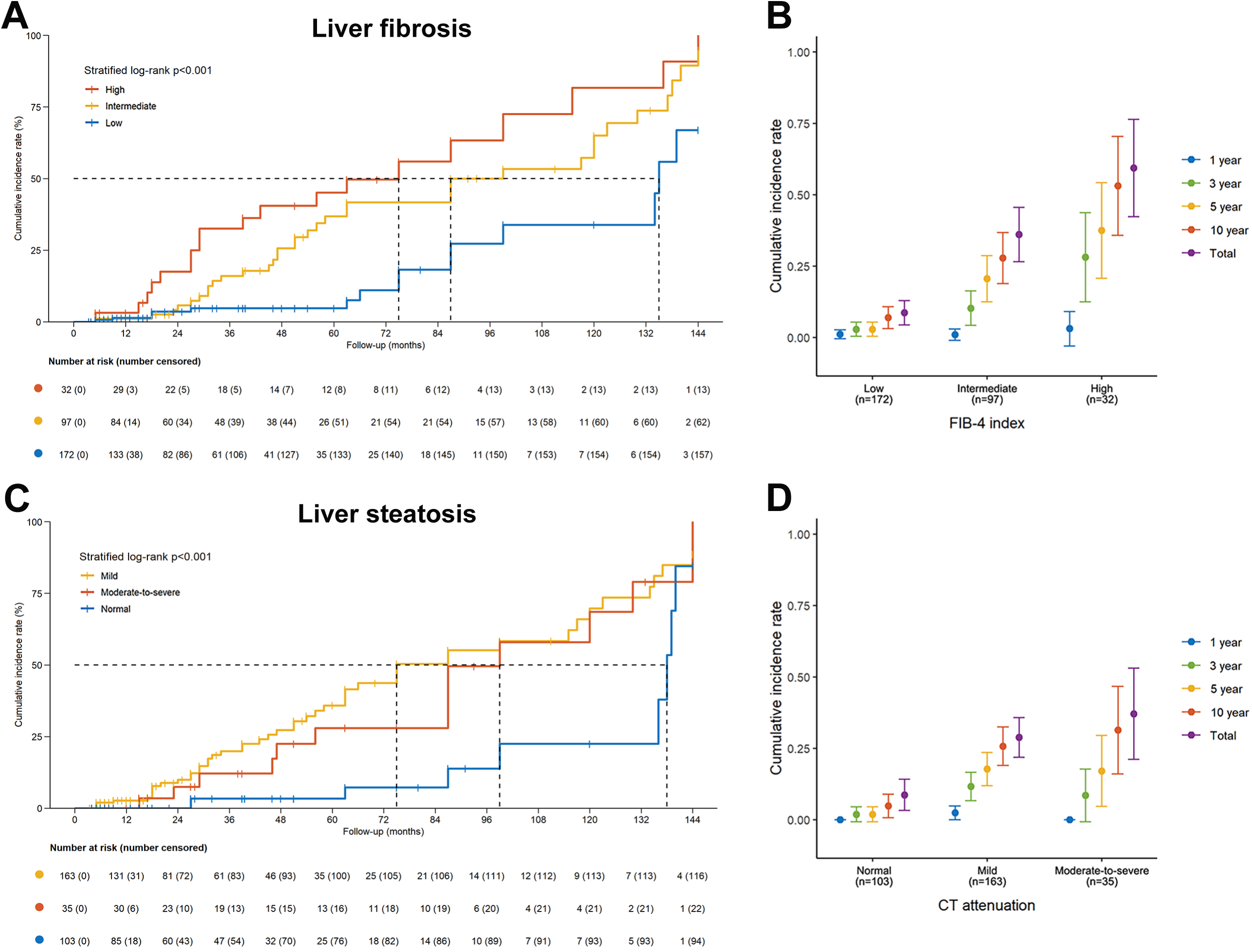
Kaplan-Meier analysis of liver fibrosis and steatosis for predicting ISR. **A)** Kaplan-Meier analysis and **B)** Cumulative incidence rate for ISR according to low, intermediate, and high FIB-4 index. **C)** Kaplan-Meier analysis and **D)** Cumulative incidence rate for ISR according to normal, mild, and moderate-to-severe steatosis. **Abbreviations:** ISR: in-stent restenosis; FIB-4: Fibrosis-4 index.

According to liver CT attenuation, patients with different liver steatosis degree were divided into three groups. The liver CT attenuation median values of the groups with normal, mild and moderate-to-severe steatosis group were 67.02 [62.5–70.82, n=103 (34.2%)], 55.20 [52.75–56.48, n=163 (54.2%)], and 38.22 [34.04–40.00, n=35 (11.6%)], respectively (P < 0.001). Kaplan-Meier survival analysis showed that ISR increased progressively with the mild and moderate-to-severe steatosis group, the median survival time were 75 and 99 months, while the median survival time of normal group was 138 months (stratified log-rank, P < 0.001) (Figure 2C). In these three steatosis groups. the respective 3, 5, and 10 year cumulative incidence rates of ISR were 1.9% (95% CI: 0.0–4.6), 1.9% (95% CI: 0.0–4.6), and 4.9% (95% CI: 0.7– 9.0) in the normal group; 11.7% (95% CI: 6.7–16.6), 17.8% (95% CI: 11.9–23.7), and 25.8% (95% CI:19.1–32.5) in the mild steatosis group; and 8.6% (95% CI: 0.0–17.8), 17.1% (95% CI: 4.7–29.6), and 31.4% (95% CI: 16.0–46.8) in the moderate-to-severe steatosis group. The 1 year cumulative incidence rates of ISR of mild steatosis group is 2.5% (95% CI: 0.1–4.8), while no patients have ISR in 1 year in normal and moderate-to-severe steatosis group (Figure 2D).

Therefore, it demonstrated that the incidence of ISR increased with the increase of patients’ FIB-4 index and steatosis degree.

### The relationship between ISR and MASLD after achieving LDL-cholesterol control standards

We selected 118 patients with LDL-cholesterol <1.8 mmol/l when near the time of occurrence of ISR, among which 78 (66.1%) had MASLD and 40 (33.9%) were non-MASLD, and 24/118 (20.3%) developed ISR. By univariate analysis, MASLD status, diabetes, FIB-4 index, liver CT attenuation and use of hypoglycemic drugs were associated with ISR (all P < 0.05) (Table 3). By multivariate analysis, FIB-4 index and liver CT attenuation were as independent factors associated with ISR [HR (95%CI): 2.72 (1.43–5.16), P = 0.002] and [HR (95%CI): 0.94 (0.88–0.99), P = 0.048). Therefore, higher FIB-4 index and liver steatosis degree (lower liver CT attenuation) could promote the occurrence of ISR even when patients achieving LDL-cholesterol control standards.

**Table 3.** Factors associated with in-stent restenosis in patients with coronary heart disease achieved LDL-cholesterol control standards.

| Factors | Univariate analysis |  | Multivariate analysis |  |
| --- | --- | --- | --- | --- |
|  | HR (95% CI) | *p* Value | HR (95% CI) | *p* Value |
| MASLD | 2.97 (1.40 ~ 6.33) | **0.005** | 2.37 (0.96 ~ 5.82) | 0.060 |
| Age (years) | 0.97 (0.93 ~ 1.01) | 0.168 |  |  |
| Female gender | 2.31 (0.85 ~ 6.22) | 0.099 |  |  |
| Smoking | 1.00 (0.44 ~ 2.27) | 0.998 |  |  |
| Alcohol | 1.05 (0.36 ~ 3.12) | 0.924 |  |  |
| Diabetes | 6.40 (1.48 ~ 27.58) | **0.013** | 3.41 (0.33 ~ 35.35) | 0.304 |
| Hypertension | 0.97 (0.33 ~ 2.87) | 0.954 |  |  |
| Systolic blood pressure (mm Hg) | 1.01 (0.99 ~ 1.04) | 0.424 |  |  |
| Diastolic blood pressure (mm Hg) | 1.02 (0.98 ~ 1.07) | 0.349 |  |  |
| Body mass index (kg/m<sup>2</sup>) | 1.02 (0.87 ~ 1.19) | 0.807 |  |  |
| Waist circumference (cm) | 1.00 (0.93 ~ 1.07) | 0.933 |  |  |
| Fasting glucose (mmol/l) | 1.05 (1.00 ~ 1.11) | **0.071** |  |  |
| Total cholesterol (mmol/l) | 1.09 (0.76 ~ 1.57) | 0.645 |  |  |
| Triglycerides (mmol/l) | 1.07 (0.84 ~ 1.36) | 0.567 |  |  |
| HDL-cholesterol (mmol/l) | 0.94 (0.18 ~ 4.78) | 0.936 |  |  |
| LDL-cholesterol (mmol/l) | 1.09 (0.66 ~ 1.79) | 0.746 |  |  |
| ApoA1 (g/L) | 0.93 (0.12 ~ 7.13) | 0.946 |  |  |
| ApoB (g/L) | 0.82 (0.11 ~ 5.99) | 0.842 |  |  |
| ApoE (mg/L) | 1.00 (0.97 ~ 1.03) | 0.995 |  |  |
| Lp-a (mg/L) | 1.00 (1.00 ~ 1.00) | 0.787 |  |  |
| Uric acid (μmol/L) | 1.00 (0.99 ~ 1.00) | 0.547 |  |  |
| Creatinine (mmol/l) | 0.99 (0.98 ~ 1.00) | 0.110 |  |  |
| Alanine aminotransferase (U/L) | 0.98 (0.95 ~ 1.00) | 0.099 |  |  |
| Platelet (10<sup>9</sup>/L) | 1.00 (0.99 ~ 1.00) | 0.326 |  |  |
| Hemoglobin A1c (%) | 1.12 (0.89 ~ 1.41) | 0.338 |  |  |
| FIB-4 index | 2.18 (1.39 ~ 3.43) | **<0.001** | 2.72 (1.43 ~ 5.16) | **0.002** |
| Liver CT attenuation (HU) | 0.95 (0.90 ~ 0.99) | **0.038** | 0.94 (0.88 ~ 0.99) | **0.048** |
| Drug use |  |  |  |  |
| Aspirin | 0.68 (0.20 ~ 2.37) | 0.550 |  |  |
| Clopidogrel | 2.23 (0.50 ~ 9.94) | 0.293 |  |  |
| Antihypertension | 0.83 (0.28 ~ 2.47) | 0.738 |  |  |
| Hypoglycemics | 2.72 (1.12 ~ 6.61) | **0.027** | 2.22 (0.62 ~ 7.98) | 0.220 |
| Stains | 0.54 (0.19 ~ 1.53) | 0.249 |  |  |

HDL, high-density lipoprotein; LDL, low-density lipoprotein; CT, computerized tomography.

### Subgroup analysis

Further confirmation of the risk stratification value of FIB-4 index for the risk of ISR was performed in subgroup analysis, as presented in Figure 3B. The result shows that in the subgroup of MASLD (no or yes), sex (male or female), age (< 65 or ≥ 65 years), BMI (< 25 or ≥ 25 kg/m^2^), drinking history (no or yes), smoking history (no or yes), hypertension (no or yes), diabetes (no or yes), and LDL (< 1.8 or ≥ 1.8 mmol/L), there were no differences in the predictive power of FIB-4 index for incidence of DES-ISR (all p for interaction > 0.05).

**Figure 3.**
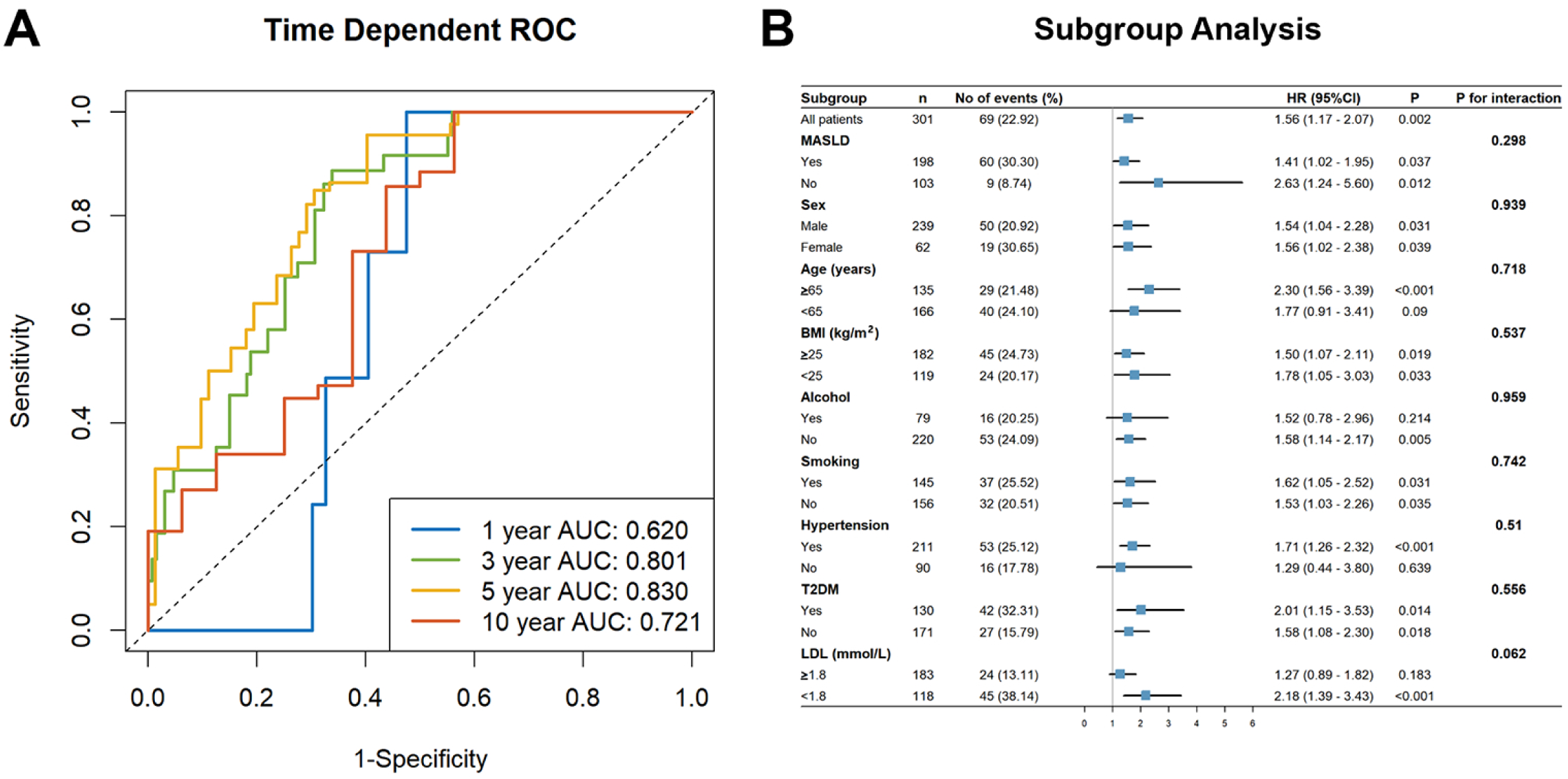
**A)** Time dependent ROC curve analysis of MASLD, FIB-4 index and age for predicting the risk of ISR in 1, 3, 5 and 10 years [AUC: 0.620 (95%CI: 0.532–0.709), AUC: 0.801 (95%CI: 0.721–0.880), AUC: 0.830 (95%CI: 0.755–0.905) and AUC: 0.721 (95%CI: 0.561–0.881) respectively]. **B)** Forest plot investigating the association between FIB-4 index and the prevalence of ISR in different subgroups. **Abbreviations:** ROC: Receiver operating characteristics curve; MASLD: metabolic dysfunction-associated steatotic liver disease; FIB-4: Fibrosis-4 index; ISR: In-stent restenosis; AUC: Area under the curve; CI: Confidence interval; BMI: body mass index; LDL-C: low-density lipoprotein cholesterol; HR: hazard ratio.

## Discussion

As the prevalence of DES-ISR increases year by year, delving into the relationship between liver steatosis and fibrosis becomes imperative. Early assessment and management of this relationship are of utmost significance for preventing cardiovascular events in patients with MASLD combined with CVD. In this retrospective observational study, 301 included CHD patients who had DES implantation underwent a median follow-up of 27 (range from 12 to 144) months to monitor for the incidence of ISR. The present study has three critical findings. First, patients with MASLD have a higher incidence of ISR than those without (30.3% vs. 8.7%). Second, MASLD status, FIB-4 index and a younger age were independent risk factors associated with ISR. Finally, LDL cholesterol reached control standard, FIB-4 index and liver CT attenuation remained independently associated with ISR. Our current study suggests there might exist a clinical correlation between MASLD and disease progression, which need to be considered for ISR risk prevention and treatment.

MASLD patients often have obesity or dysglycemia and dyslipidemia, which is considered to be manifestations of metabolic syndrome, which may lead to intimal hyperplasia and atherosclerosis and accelerate the development of ISR [10]. By using a univariate cox proportional hazard model, we confirmed that MASLD [HR (95%CI): 3.49 (1.73–7.03), P < 0.001], fasting glucose [HR (95%CI): 1.06 (1.01– 1.11), P = 0.035], total cholesterol [HR (95%CI): 1.20 (1.02–1.42), P = 0.031], triglycerides [HR (95%CI): 1.48 (1.18–1.87), P < 0.001] were all associated with ISR. Additionally, LDL-cholesterol [HR (95%CI): 1.26 (0.99–1.60), P = 0.059] and Hemoglobin A1c [HR (95%CI): 1.20 (1.00–1.45), P = 0.050] showed a relative relation with ISR with borderline statistical significance. MASLD is characterized by excessive triglycerides, cholesterol, and free fatty acids, which provide abundant oxidized lipid raw materials and move through the neointimal promoting the formation of neo-atherosclerosis·[11,12]. Moreover, its induced lipotoxicity further aggravate insulin resistance and induce systemic inflammatory response by releasing inflammatory factors and C-reactive protein, which in turn causes vascular endothelial dysfunction [13]. Many previous studies in both humans and animal models of MASLD that have implicated increases in hepatic diacylglycerol content leading to activation of novel protein kinase Cɛ resulting in decreased insulin signaling [14]. Insulin resistance leads to endothelial cell dysfunction by reducing endothelial cell synthesis of nitric oxide (NO) and increasing the release of procoagulant factors that promote platelet aggregation, finally leads to the formation of neo-atherosclerosis [15,16].

Further in multivariate analysis we found MASLD, higher FIB-4 index and a younger age were independent risk factors associated with ISR, and had relatively good predictive value for ISR (AUC of 1, 3, 5 and 10 years prediction were respectively 0.620, 0.801, 0.830 and 0.72). The incidence rate of ISR increased progressively with the low, intermediate and high FIB-4 index group. Although the apparent mechanisms responsible for the relationship between liver fibrosis and atherosclerosis have not been fully elucidated, one possible reason is endothelial dysfunction triggered by persistent chronic inflammation and oxidative stress [17,18]. A previous study showed that liver fibrosis promoted angiogenesis and vasoconstriction, contributing to inflammation and fibrosis in vascular tissues, and might cause intimal hyperplasia [19]. Both ISR and liver fibrosis are closely associated with inflammatory response. In ISR, endothelial damage activates inflammatory response, leading to leukocyte aggregation and cytokine release (such as interleukin-1 (IL-1), interleukin-6 (IL-6), and tumor necrosis factor-alpha (TNF-α), etc.), which in turn promotes smooth muscle cell proliferation and migration, leading to neointimal formation and restenosis. Similarly, liver fibrosis is also caused by chronic inflammation. Hepatic stellate cells are activated and transformed into profibrotic cells, leading to liver fibrosis [20]. In addition, because of the proliferation of collagen fibers, which is one of the vital matrices for vessel walls and essential elements in liver fibrosis, there is a possibility that similar cell signaling pathways and molecular mechanisms in the arterial wall could be expected to increase coronary arterial stiffness [21,22]. Adiponectin has been demonstrated to have an anti-fibrotic action in the liver by blocking the activation of hepatic stellate cell [23]. And by modulating endothelial function and inflammatory responses, adiponectin can also offer a protective mechanism against cardiovascular pathology, so adiponectin levels may signify a risk factor linking liver fibrosis and atherosclerosis [24].

The increased susceptibility of younger patients to ISR may be attributed to their stronger immune systems. This heightened immune activity may result in a more pronounced inflammatory response following stent implantation. Consequently, the proliferation and migration of vascular smooth muscle cells are enhanced, ultimately promoting intimal hyperplasia. [25]. Alternatively, younger patients may harbor genetic susceptibility factors associated with ISR, while this hypothesis requires further investigation [26].

According to the latest guidelines for the management of chronic coronary syndromes, LDL cholesterol in patients with CVD should be reduced to the lowest possible level. In our retrospective study, LDL-cholesterol control standard was defined as <1.8 mmol/L and decreased by >50% from baseline [27]. Although LDL cholesterol was controlled <1.8 mmol/L by lipid lowering therapy, there were still 20.3% patients developed ISR. By using univariate analysis, diabetes [HR (95%CI): 6.40 (1.48 ∼ 27.58), P=0.013] was still related to ISR, which was consistent with previous studies. After adjusting for confounding factors, the degree of liver fibrosis and hepatic steatosis are independent risk factors for ISR. In patients with MASLD/MASH, macrophages in the liver secrete various pro-inflammatory cytokines, which may induce endothelial dysfunction, promote neointimal formation, and facilitate the progression of atherosclerotic plaques. These distant-acting inflammatory factors may be independent of lipid levels and increase the risk of ISR [28,29]. Accompanying hepatic steatosis, the increase in visceral fat, particularly epicardial adipose tissue, can secrete various inflammatory and chemotactic factors via paracrine mechanisms [30]. These factors directly act on the coronary arteries, leading to local inflammation and the formation of neointima and atherosclerosis [31,32].

In contrast, angina pectoris or myocardial infarction may promote hepatic fibrosis in patients with MASLD/MASH, but it is evident that both conditions can alter systemic homeostasis and are closely linked through immune-inflammatory responses [33].

An animal studies have demonstrated that trans fat-induced MASLD increases the accumulation of sphingolipids in the liver and promotes the secretion of very-low-density lipoprotein (VLDL) which are closely associated with the development of atherosclerosis [34]. Meanwhile, a large-scale study showed that Lipoprotein(a) is an independent predictor for long-term ISR. The surface of Lp(a) is coated with phospholipids and free cholesterol, and may also contain oxidized phospholipids (OxPL) which may be possible factors for ISR [35].

### Limitations

The present study has several limitations, mainly due to its retrospective design. First, the diagnosis of MASLD was based on ultrasound and computerized tomography findings, which might be an insufficient method for evaluating the degree of liver steatosis. As a result, mild steatosis cases may have been underdiagnosed. Second, each score was calculated based on the screening time, and the related follow-up data were unavailable. Some patients may develop liver fibrosis during follow-up, resulting in misclassifications. Third, we could not evaluate the relationship between these liver fibrosis scores and the severity of liver fibrosis because of the lack of liver fibrosis transient elastography. Furthermore, no data on liver biopsy evaluation were available, which is one of the gold-standard methods for evaluating the severity of liver fibrosis. Moreover, the precise mechanism underlying the difference between baseline liver fibrosis and ISR has not been fully elucidated. Thus, further extensive investigation is warranted.

## Conclusions

MASLD and FIB-4 index are associated with the risk of ISR. The occurrence of ISR increased progressively with the FIB-4 index. After LDL cholesterol achieving control standards, FIB-4 index is still an independent risk factor of ISR. CHD patients need to pay more attention to the screening and treatment of MASLD and related liver fibrosis after DES implantation.

## Abbreviations

AUC: areas under the receiver operator characteristic curve
ALT: alanine aminotransferase
AST: aspartate transaminase
BMI: body mass index
CVD: cardiovascular disease
CT: computerized tomography
CI: confidence interval
CHD: coronary heart disease
DES: drug-eluting stents
FIB-4: fibrosis-4 index
HR: hazard ratio
HDL: high-density lipoprotein cholesterol
HU: hounsfield units
ISR: in-stent restenosis
IQR: inter quartile range
IL: interleukin
IVUS: intravenous ultrasound
LDL: low-density lipoprotein cholesterol
MASH: metabolic dysfunction-associated steatohepatitis
MASLD: metabolic dysfunction-associated steatotic liver disease
NAFLD: non-alcoholic fatty liver disease
OxPL: oxidized phospholipid
PCI: percutaneous coronary intervention
ROC: receiver operating characteristic
SD: standard deviation
TNF-α: tumor necrosis factor-alpha
VLDL: very-low-density lipoprotein

## Author Contributions

Conception and design: Z.B and Y.J; administrative support: Z.B and Y.J; provision of study materials or patients: L.J and Z.X; collection and assembly of data: L.J and Z.X; data analysis and interpretation: L.J, L.L, T.L and S.C; manuscript writing: all authors. All authors have read and agreed to the published version of the manuscript.

## Funding

This work was supported by the National Natural Science Foundation of China [grant numbers 82370587, 82400663, 81870404, 82100648]; the Natural Science Foundation of Guangdong Province, China [grant number 2022A1515012369]; and the China Postdoctoral Science Foundation [grant number 2020M683128].

## Data Availability Statement

Data described in the manuscript, code book, and analytic code will be made publicly and freely available without restriction.

## Acknowledgments

We are grateful to Aihua Lin in the School of Public Health at Sun Yat-sen University for her assistance in the statistical analysis of this study. Additionally, we would like to thank Kimi (https://kimi.moonshot.cn/) for English language editing.

## Conflicts of Interest

The authors declare no conflict of interest.

## Notes

### Competing Interest Statement

The authors have declared no competing interest.

### Author Declarations

This retrospective observational cohort study was conducted utilizing data from the electronic specialize disease of Cardiovascular disease Database (CDD) managed by the at the Fatty Liver Disease Center and Cardiology of the First Affiliated Hospital of Sun Yat-sen University (Guangzhou, China). CDD is a comprehensive electronic database collecting demographic data, laboratory test information, diagnoses, treatment prescriptions, follow-up status, as well as through capturing inpatient data of the First Affiliated Hospital, Sun Yat-sen university in Guangzhou, representing the largest hospital in southern China. The study was approved by the Ethics Committee of the First Affiliated Hospital, Sun Yat-sen university {[2014]112}. All study participants provided signed informed consent.

